# Impact of COVID-19 on recorded blood pressure screening and hypertension management in England: An analysis of monthly changes in Quality and Outcomes Framework indicators in OpenSAFELY

**DOI:** 10.1101/2023.07.20.23292883

**Authors:** Milan Wiedemann, Victoria Speed, Christine Cunningham, Rose Higgins, Helen J Curtis, Colm Andrews, Louis Fisher, Lisa E M Hopcroft, Christopher T Rentsch, Viyaasan Mahalingasivam, Laurie Tomlinson, Caroline E Morton, Miriam Samuel, Amelia C A Green, Christopher Wood, Andrew Brown, Jon Massey, Caroline Walters, Rebecca Smith, Peter Inglesby, Dave Evans, Steve Maude, Iain Dillingham, Alex J Walker, Jessica Morley, Amir Mehrkar, Sebastian C J Bacon, Christopher Bates, Jonathan Cockburn, John Parry, Frank Hester, Sam Harper, Richard J McManus, Ben Goldacre, Brian MacKenna

## Abstract

**Background:** Cardiovascular disease management in primary care in England was disrupted during the COVID-19 pandemic.

**Objective:** To describe the impact of the COVID-19 pandemic on blood pressure screening and hypertension management, based upon a national quality of care scheme (Quality and Outcomes Framework, QOF) across key demographic, regional, and clinical subgroups. To this end, we translated complex clinical quality of care schemes from text descriptions into reusable analytic code.

**Methods:** With the approval of NHS England, a population based cohort study was conducted on 25.2 million patient records in situ using OpenSAFELY-TPP. We included all NHS patients registered at general practices using TPP software between March 2019 and March 2023. Individuals that were eligible for blood pressure screening and with a diagnosis of hypertension were identified according to the QOF 2021/22 business rules. We examined monthly changes in recorded blood pressure screening in the preceding 5 years in patients aged ≥ 45, recorded hypertension prevalence, and the recorded percentage of patients treated to target (i.e., ≤ 140/90 mmHg for patients ≤ 79 years and ≤ 150/90 mmHg for patients ≥ 80 years) in the preceding 12 months, within demographic, regional, and clinical subgroups as well as the variation across practices.

**Results:** The overall percentage of patients aged ≥ 45 who had blood pressure screening recorded in the preceding 5 years decreased from 90% in March 2019 to 85% in March 2023. Recorded hypertension prevalence was relatively stable at 15% throughout the study period. The percentage of patients with a record of hypertension treated to target in the preceding 12 months reduced from a maximum of 71% in March 2020 to a minimum of 47% in February 2021 in patients aged ≤ 79 years, and from 85% in March 2020 to a minimum of 58% in February 2021 in patients aged ≥ 80 years before recovering. Blood pressure screening rates in the preceding 5 years remained stable in older age groups, patients with a record of learning disability, or care home status.

**Conclusions:** There was substantial disruption to hypertension management QOF indicators during the pandemic, which can likely be attributed to a general reduction of blood pressure measurement including screening. OpenSAFELY can be used to continuously monitor monthly changes in national quality of care schemes to identify changes in key clinical subgroups early and support prioritisation of recovery from disrupted care caused by COVID-19.

## Introduction

The COVID-19 pandemic disrupted healthcare services globally [1]. Cardiovascular disease (CVD) management in primary care in England was impacted [2–4] with an estimated 2,175 non-COVID excess deaths attributed to hypertensive diseases in between March 2020 and December 2021 [5]. Cardiovascular disease is associated with a higher risk of morbidity and mortality from COVID-19, emphasising the importance of maintaining good routine care [6,7]. High blood pressure is the leading risk factor for cardiovascular disease [8] and one of the top three risk factors for global disease burden [9]. Since 1994 there has been improvement in the management of high blood pressure in England [10] and a reduction of the negative impact of social deprivation on blood pressure management [11]. Delayed management of hypertension is associated with worse clinical outcomes, for example stroke [12]. Recent results from annual national audits of England’s population on cardiovascular disease and NHS Digital have also suggested that blood pressure management was disrupted by the pandemic [13,14].

In 2004 the Quality and Outcomes Framework (QOF) was introduced in England as one of the largest initiatives worldwide to improve the quality of care in general practice. General practitioners (GPs) and their staff are measured on indicators of good clinical care and receive financial incentives based on their achievement of certain thresholds [15,16]. In order to monitor the indicators and thresholds, NHS Digital publishes text descriptions of analytic rules and logic, commonly referred to as ‘*business rules’* which are taken by software providers and implemented in GP electronic health records systems [17]. GPs can review their delivery of care against these rules and indicators for their practice throughout the financial year, however, national data for all practices is only available annually. At the end of every NHS financial year on March 31st, NHS Digital calculates each practice’s achievement against set thresholds for individual indicators. Between 1st April 2020 and 31st March 2023, amendments were made to QOF and some preventative indicators were suspended, including hypertension management, to support the COVID-19 response and support roll out of the national COVID-19 vaccination program [18,19].

OpenSAFELY is a secure analytics platform for electronic patient records built by our group on behalf of NHS England to deliver urgent academic [6,20,21] and operational research [2,22,23] during the pandemic. Using OpenSAFELY-TPP, we therefore aimed to describe trends and variation in these indicators before and during the COVID-19 pandemic and assess recovery of the indicators to pre-pandemic levels across key clinical and demographic subgroups.

## Methods

### Data source

Primary care records managed by the GP software provider TPP were accessed through OpenSAFELY (https://opensafely.org). OpenSAFELY is a secure analytics platform for electronic patient records built by our group with the approval of NHS England to deliver urgent academic [6] and operational NHS service research [22,23] on the direct and indirect impacts of the pandemic.

OpenSAFELY provides a secure software interface allowing the analysis of pseudonymized primary care patient records from England. in near real-time within the EHR vendor’s highly secure data centre, avoiding the need for large volumes of potentially disclosive pseudonymized patient data to be transferred off-site. The dataset analysed within OpenSAFELY is based on 25 million people currently registered with GP surgeries using TPP SystmOne software. It includes pseudonymized data such as coded diagnoses, medications and physiological parameters. No free text data is included. Further details on our information governance and ethics can be found in the Appendix.

### Study design and population

We conducted a retrospective cohort study from March 2019 to March 2023 using primary care EHR data from all GP practices in England supplied by the EHR vendor TPP, a cohort that is broadly representative of the population in England [25]. Following the QOF business rules, we included all patients who were alive and registered with an OpenSAFELY-TPP practice for the QOF hypertension prevalence and management indicators (HYP001, HYP003, HYP007). For the QOF blood pressure screening indicator (BP002) we included only those aged ≥ 45.

### Implementation of QOF business rules in analytic code

QOF indicators for blood pressure screening in the preceding five years (BP002), hypertension register (HYP001) and hypertension management in the preceding 12 months (HYP003, HYP007) were specified in analytic code replicating the QOF business rules for 2021/22 [Version 46, 17] using the OpenSAFELY framework (Table 1). All QOF indicators are formed by specifying rules and logic which determine aggregate counts of patients. Percentages are then calculated using numerator and denominator pairs. In addition to QOF indicators we also applied the same clinical rules for blood pressure screening (BP002) to all patients with a record of an unresolved hypertension diagnosis using a 12 months lookback period to match the timeframe used in the hypertension management indicators (HYP003 and HYP007).

**Table 1.** Descriptions of the QOF indicators for blood pressure (BP) and hypertension (HYP).

| Indicator | Domain / Category | Indicator description | Population of interest |
| --- | --- | --- | --- |
| BP002 | Public health | The percentage of patients aged 45 or over who have a record of blood pressure **in the preceding 5 years**. | All registered patients aged $\geq 45$ |
| HYP001* | Clinical / Records | The contractor establishes and maintains a register of patients with established hypertension. | All registered patients |
| HYP003<sup>†</sup> | Clinical / Ongoing management | The percentage of patients aged 79 years or under, with hypertension, in whom the last blood pressure reading (measured **in the preceding 12 months**) is 140/90 mmHg or less. | Hypertension register (HYP001*) |
| HYP007<sup>†</sup> | Clinical / Ongoing management | The percentage of patients aged 80 years or over, with hypertension, in whom the last blood pressure reading (measured **in the preceding 12 months**) is 150/90 mmHg or less. | Hypertension register (HYP001*) |

*Note.* \* Indicator HYP001 refers to the hypertension register (HYP\_REG) which is defined as 'Patients with an unresolved diagnosis of hypertension'. <sup>†</sup> For the purpose of this analysis, this is the treatment target. The population that meets the blood pressure target will be described as *treated to target*. More details on the specific select and exclusion rules and the codelists are available in the Supplementary Appendix (Tables B1 - B4 and C1).

Higher indicator percentages represent a higher percentage of patients receiving indicated clinical care. Patients can be excluded from the denominator according to QOF rules, such as those who declined treatment (for more details see Appendix A). For the two hypertension control indicators (HYP003 and HYP007), patients with hypertension who did not have their blood pressure recorded in the last year are counted as not being treated to target as per the QOF business rules.

Data were analysed for each month between 1st March 2019 and 31st March 2023 covering five financial years. Each monthly cohort replicated the yearly reporting of each QOF business rules. Thus, the data presented for each March in this study aligns with the reporting period of the corresponding annual QOF reports published by NHS Digital.

### Monthly changes in QOF indicators across demographic, regional, and clinical subgroups

Trends and variation in QOF indicators were reported across demographic (10 year age bands, sex, ethnicity in 5 and 16 categories), regional (practice level deciles, Indices of Multiple Deprivation quintiles derived from patient’s postcode at lower super output area, region), and key clinical subgroups (record of learning disability and care home status) highlighted in the NHS long term plan as priority groups [26].

### Software and reproducibility

Data management and analyses were performed using the OpenSAFELY software libraries using Python (Version 3.8.10) and R (Version 4.0.2). We used the R packages *ggplot2* [27] and *gt* [28] to visualise data and create tables. Code replicating the QOF business rules are available at https://github.com/opensafely/hypertension-sro and https://github.com/opensafely/blood-pressure-sro alongside all analytic code and codelists. The GitHub repository https://github.com/opensafely/qof-utilities contains reusable code developed for implementing QOF rules in OpenSAFELY.

### Patient and public involvement

For transparency purposes we have developed a public website (https://opensafely.org/) which provides a detailed description of the platform in language suitable for a lay audience; we have participated in two citizen juries exploring public trust in OpenSAFELY [29]. To ensure the patient voice is represented, we are working closely with appropriate medical research charities however there was no patient or public involvement in this specific research question.

## Results

### Calculating monthly trends in QOF indicators

Detailed demographic characteristics of all patients considered for the blood pressure and hypertension indicators during the reporting period of the NHS financial year 21/22 are presented in Table 2.

**Table 2.** Cohort description for patients included in the blood pressure and hypertension QOF indicators in OpenSAFELY-TPP during the NHS financial year 21/22. All counts were rounded to the nearest 10.

|  | BP002 (Age >= 45) |  |  | HYP001 (Total population) |  |  | HYP003 (Age <= 79) |  |  | HYP007 (Age >= 80) |  |  |
| --- | --- | --- | --- | --- | --- | --- | --- | --- | --- | --- | --- | --- |
|  | Numerator | Denominator | Receiving indicated care | Register | List size | Prevalence | Numerator | Denominator | Receiving indicated care | Numerator | Denominator | Receiving indicated care |
| **Population** | 9,587,610 | 11,195,670 | 85.6% | 3,672,870 | 25,287,730 | 14.5% | 1,607,520 | 2,664,840 | 60.3% | 554,240 | 764,960 | 72.5% |
| **Sex** |  |  |  |  |  |  |  |  |  |  |  |  |
| Female | 5,115,540 | 5,736,940 | 89.2% | 1,848,910 | 12,621,240 | 14.7% | 782,460 | 1,268,720 | 61.7% | 326,560 | 463,200 | 70.5% |
| Male | 4,472,070 | 5,458,730 | 82.0% | 1,823,960 | 12,666,500 | 14.4% | 825,060 | 1,396,110 | 59.1% | 227,680 | 301,760 | 75.5% |
| **Age band** |  |  |  |  |  |  |  |  |  |  |  |  |
| 0-19 | - | - | - | 2,180 | 5,547,410 | 0.04% | 1,010 | 1,780 | 56.7% | - | - | - |
| 20-29 | - | - | - | 11,400 | 3,139,340 | 0.4% | 4,790 | 9,120 | 52.5% | - | - | - |
| 30-39 | - | - | - | 60,680 | 3,629,760 | 1.7% | 25,760 | 50,520 | 51.0% | - | - | - |
| 40-49 | 1,124,490 | 1,548,870 | 72.6% | 220,760 | 3,246,940 | 6.8% | 99,700 | 191,390 | 52.1% | - | - | - |
| 50-59 | 2,751,360 | 3,415,030 | 80.6% | 615,040 | 3,458,290 | 17.8% | 308,270 | 555,420 | 55.5% | - | - | - |
| 60-69 | 2,407,340 | 2,752,690 | 87.5% | 887,520 | 2,774,540 | 32.0% | 506,800 | 828,700 | 61.2% | - | - | - |
| 70-79 | 2,079,410 | 2,215,190 | 93.9% | 1,079,680 | 2,224,430 | 48.5% | 661,200 | 1,027,890 | 64.3% | - | - | - |
| 80+ | 1,225,010 | 1,263,890 | 96.9% | 795,620 | 1,267,020 | 62.8% | - | - | - | 554,240 | 764,960 | 72.5% |
| **Ethnicity** |  |  |  |  |  |  |  |  |  |  |  |  |
| **Asian or Asian British** |  |  |  |  |  |  |  |  |  |  |  |  |
| Any other Asian background | 106,280 | 126,810 | 83.8% | 39,230 | 412,650 | 9.5% | 21,570 | 33,220 | 64.9% | 2,250 | 3,080 | 73.1% |
| Bangladeshi | 27,800 | 30,490 | 91.2% | 11,860 | 124,500 | 9.5% | 6,900 | 10,060 | 68.6% | 570 | 820 | 69.5% |
| Indian | 209,490 | 239,700 | 87.4% | 91,540 | 710,370 | 12.9% | 47,230 | 74,360 | 63.5% | 8,100 | 11,090 | 73.0% |
| Pakistani | 123,280 | 136,790 | 90.1% | 48,560 | 513,040 | 9.5% | 25,980 | 40,040 | 64.9% | 3,530 | 4,900 | 72.0% |
| **Black or Black British** |  |  |  |  |  |  |  |  |  |  |  |  |
| African | 90,470 | 108,040 | 83.7% | 40,280 | 385,090 | 10.5% | 18,710 | 34,270 | 54.6% | 1,000 | 1,620 | 61.7% |
| Any other Black background | 29,650 | 34,290 | 86.5% | 11,700 | 103,910 | 11.3% | 5,340 | 9,760 | 54.7% | 570 | 840 | 67.7% |
| Caribbean | 55,000 | 61,340 | 89.7% | 27,660 | 115,680 | 23.9% | 11,410 | 19,970 | 57.1% | 4,160 | 5,900 | 70.5% |
| **Mixed** |  |  |  |  |  |  |  |  |  |  |  |  |
| Any other mixed background | 22,820 | 28,270 | 80.7% | 7,480 | 141,140 | 5.3% | 3,600 | 6,150 | 58.5% | 450 | 630 | 71.4% |
| White and Asian | 12,650 | 15,320 | 82.6% | 3,970 | 81,280 | 4.9% | 1,980 | 3,270 | 60.6% | 240 | 340 | 70.6% |
| White and Black African | 13,620 | 16,600 | 82.1% | 5,660 | 70,890 | 8.0% | 2,590 | 4,770 | 54.3% | 150 | 250 | 60.0% |
| White and Black Caribbean | 15,690 | 17,990 | 87.2% | 6,710 | 84,870 | 8.0% | 2,840 | 5,160 | 55.0% | 690 | 970 | 71.1% |
| **Other Ethnic Groups** |  |  |  |  |  |  |  |  |  |  |  |  |
| Any other ethnic group | 69,430 | 89,460 | 77.6% | 21,420 | 330,160 | 6.5% | 10,320 | 17,140 | 60.2% | 1,610 | 2,210 | 72.9% |
| Chinese | 31,660 | 43,510 | 72.8% | 9,030 | 182,960 | 5.0% | 4,520 | 7,100 | 63.7% | 720 | 1,030 | 69.9% |
| **White** |  |  |  |  |  |  |  |  |  |  |  |  |
| Any other White background | 589,110 | 755,220 | 78.0% | 220,030 | 2,320,120 | 9.5% | 92,240 | 163,970 | 56.3% | 26,690 | 37,150 | 71.84% |
| British | 6,951,180 | 7,855,620 | 88.5% | 2,607,310 | 14,450,100 | 18.0% | 1,151,190 | 1,881,440 | 61.2% | 409,870 | 559,430 | 73.27% |
| Irish | 58,040 | 66,660 | 87.1% | 22,080 | 112,460 | 19.6% | 8,790 | 14,600 | 60.2% | 4,360 | 6,020 | 72.43% |
| **(Missing)** | 1,181,420 | 1,569,530 | 75.3% | 498,350 | 5,148,500 | 9.7% | 192,300 | 339,570 | 56.6% | 89,300 | 128,680 | 69.40% |
|  | Numerator | Denominator | Receiving indicated care | Register | List size | Prevalence | Numerator | Denominator | Receiving indicated care | Numerator | Denominator | Receiving indicated care |
| **IMD** |  |  |  |  |  |  |  |  |  |  |  |  |
| 1 - Most deprived | 1,541,010 | 1,793,920 | 85.9% | 634,660 | 4,993,420 | 12.7% | 298,170 | 486,270 | 61.3% | 74,240 | 102,320 | 72.6% |
| 2 | 1,728,400 | 2,015,760 | 85.7% | 688,070 | 4,885,930 | 14.1% | 307,680 | 509,890 | 60.3% | 94,540 | 130,790 | 72.3% |
| 3 | 2,078,700 | 2,425,360 | 85.7% | 799,060 | 5,203,780 | 15.4% | 344,130 | 573,440 | 60.0% | 126,130 | 173,430 | 72.7% |
| 4 | 2,062,080 | 2,410,990 | 85.5% | 769,580 | 4,892,080 | 15.7% | 327,280 | 545,930 | 60.0% | 126,760 | 175,140 | 72.4% |
| 5 - Least deprived | 1,977,820 | 2,313,780 | 85.5% | 709,070 | 4,503,140 | 15.8% | 297,250 | 496,010 | 59.9% | 123,440 | 170,860 | 72.3% |
| (Missing) | 199,590 | 235,860 | 84.6% | 72,440 | 809,380 | 9.0% | 33,000 | 53,300 | 61.9% | 9,140 | 12,420 | 73.6% |
| **Region** |  |  |  |  |  |  |  |  |  |  |  |  |
| East | 2,190,210 | 2,552,650 | 85.8% | 828,930 | 5,785,610 | 14.3% | 349,120 | 596,850 | 58.5% | 126,920 | 178,030 | 71.3% |
| East Midlands | 1,689,470 | 1,966,690 | 85.9% | 658,520 | 4,391,220 | 15.0% | 295,020 | 481,870 | 61.2% | 97,480 | 132,650 | 73.5% |
| London | 467,680 | 581,050 | 80.5% | 174,910 | 1,798,910 | 9.7% | 87,120 | 138,270 | 63.0% | 19,930 | 27,610 | 72.2% |
| North East | 444,680 | 516,940 | 86.0% | 175,280 | 1,176,200 | 14.9% | 81,040 | 129,640 | 62.5% | 26,460 | 35,520 | 74.5% |
| North West | 896,960 | 1,034,710 | 86.7% | 355,940 | 2,185,660 | 16.3% | 172,190 | 263,810 | 65.3% | 51,580 | 69,400 | 74.3% |
| South East | 657,640 | 775,260 | 84.8% | 238,870 | 1,647,190 | 14.5% | 91,050 | 167,610 | 54.3% | 37,710 | 55,340 | 68.1% |
| South West | 1,484,430 | 1,733,970 | 85.6% | 547,440 | 3,508,660 | 15.6% | 219,820 | 375,880 | 58.5% | 94,190 | 129,810 | 72.6% |
| West Midlands | 355,330 | 412,660 | 86.1% | 144,560 | 1,041,080 | 13.9% | 61,510 | 108,300 | 56.8% | 20,180 | 28,890 | 69.9% |
| Yorkshire and The Humber | 1,374,980 | 1,593,520 | 86.3% | 538,010 | 3,666,360 | 14.7% | 246,450 | 397,970 | 61.9% | 78,570 | 106,420 | 73.8% |
| **Care home status** |  |  |  |  |  |  |  |  |  |  |  |  |
| No | 9,443,510 | 11,049,600 | 85.5% | 3,617,810 | 25,175,090 | 14.37% | 1,599,120 | 2,652,850 | 60.3% | 526,350 | 727,500 | 72.4% |
| Yes | 144,100 | 146,060 | 98.7% | 55,060 | 112,640 | 48.88% | 8,390 | 11,980 | 70.0% | 27,890 | 37,460 | 74.5% |
| **Record or learning disability** |  |  |  |  |  |  |  |  |  |  |  |  |
| No | 9,542,000 | 11,149,060 | 85.6% | 3,658,810 | 25,144,720 | 14.55% | 1,598,740 | 2,652,360 | 60.3% | 553,820 | 764,410 | 72.5% |
| Yes | 45,610 | 46,610 | 97.6% | 14,060 | 143,010 | 9.83% | 8,780 | 12,480 | 70.4% | 420 | 550 | 76.4% |

**Notes.** BP002 = the percentage of patients aged ≥ 45 with blood pressure screening in the preceding 5 years; HYP001 = hypertension register; HYP003 and HYP007 = the percentage of patients aged ≤ 79 years and ≥ 80 respectively diagnosed with hypertension and treated to target in the preceding 12 months.

### Changes in blood pressure screening and hypertension management rates in the total population

In the total population, the percentage of patients aged ≥ 45 (BP002) with recorded blood pressure in the preceding 5 years decreased steadily from its maximum of 90.4% in March 2019 to a minimum of 87.2% in March 2023 (Figure 1A).

**Figure 1.**
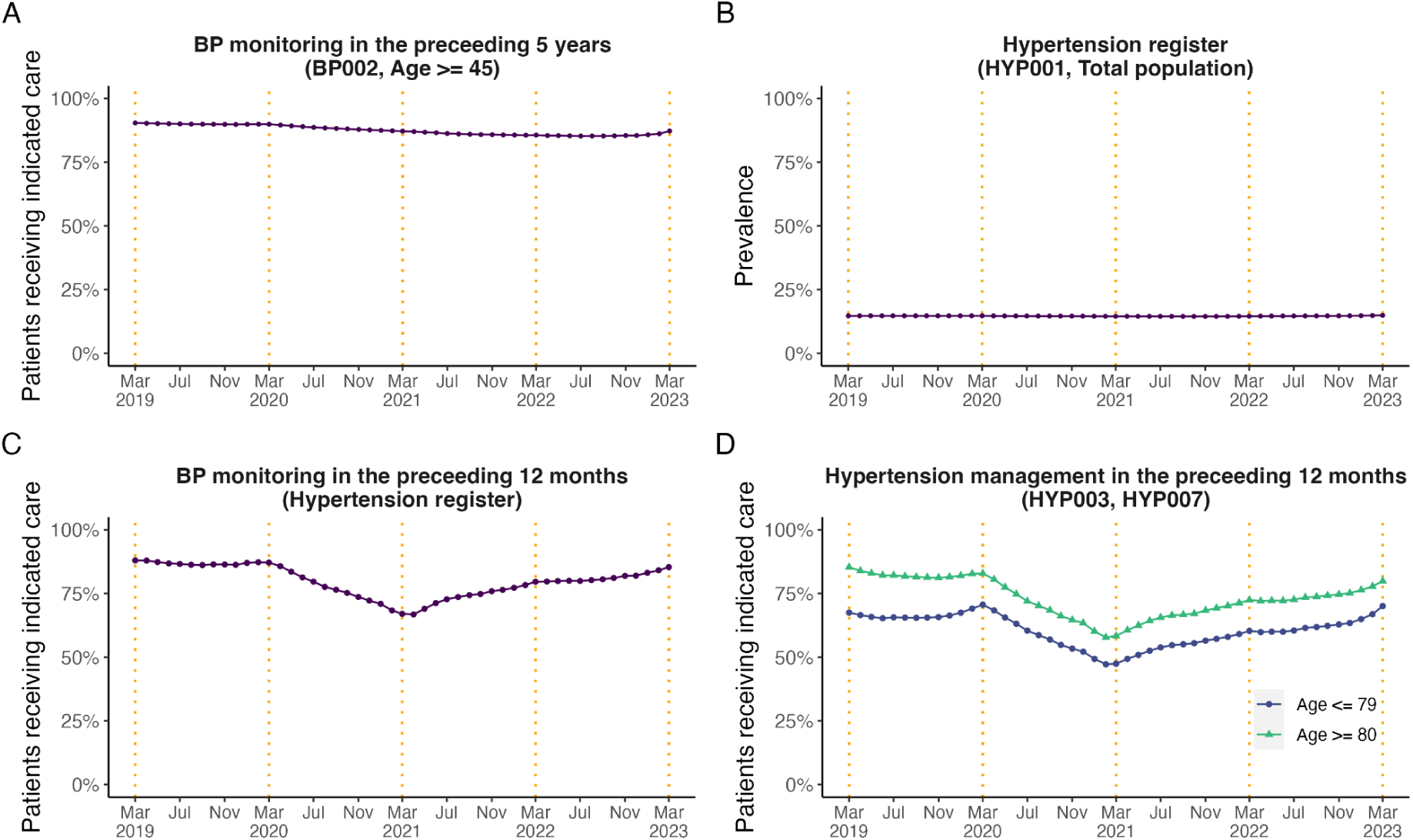
Monthly trends from March 2019 to March 2023 in (A) the percentage of patients aged ≥ 45 with blood pressure screening in the preceding 5 years (BP002); (B) the hypertension register (HYP001); (C) the percentage of patients diagnosed with hypertension and recorded blood pressure in the preceding 12 months. Demographic, regional, and clinical subgroups for panel C are presented in the Appendix (Figure D1); (D) the percentage of patients diagnosed with hypertension and treated to target (HYP003 and HYP007) in the preceding 12 months. BP = Blood pressure; HYP = Hypertension. The end of the NHS financial years (March) are highlighted with orange dashed vertical lines. Counts of patients in the numerator and denominator pair are presented in the Appendix (Figures C1 and C2).

Recorded hypertension prevalence (HYP001) was relatively stable throughout the entire study period (14.7% in March 2019 to 14.9% in March 2023, Figure 1B). Blood pressure screening in the preceding 12 months in patients identified as having hypertension decreased from its peak of 88% in March 2019 to its lowest value of 68% in April 2021 and subsequently improved steadily to 85% in March 2023.

Of those aged ≤ 79 identified as having hypertension, the percentage of patients with blood pressure treated to target (HYP003) varied from 67.5% in March 2019 to 70.1% in March 2023, with a peak of 70.6% in March 2020 and a lowest value of 47.2% in February 2021 (Figure 1C). Of those aged ≥ 80 years the percentage of patients with blood pressure treated to target (HYP007) reduced from 85.3% in March 2019 (the peak value) to 79.9% in March 2023, with a lowest value of 57.8% in February 2021 (Figure 1D). For both hypertension management indicators (HYP003 and HYP007) as well as blood pressure screening in patients identified as having hypertension, the results indicated a steady improvement between March 2021 to March 2023.

By March 2023, the differences compared to March 2019 were a 3.1% decrease (90.4% to 87.2%) in patients aged ≥ 45 (BP002) with recorded blood pressure in the preceding 5 years, a 0.2% increase (14.7% to 14.9%) in recorded hypertension prevalence (HYP001), a 2.6% increase (67.5% to 70.1%) and a 5.4% decrease (85.3% to 79.9%) in patients identified as having hypertension with blood pressure treated to target aged ≤ 79 and ≥ 80 years respectively (HYP003 and HYP007). Counts of patients in the numerator and denominator are presented in the Appendix (Figure C1).

### Changes in blood pressure screening and hypertension management rates in demographic, regional, and clinical subgroups

#### Subgroups for blood pressure screening in the preceding 5 years in patients aged ≥ 45 (BP002)

Preexisting differences in blood pressure screening rates between younger and older age groups increased over the study period, with a reduction in screening observed in younger adults (e.g., for age category 45 to 49: 81.9% in March 2019 to 74.9% in March 2023) but not older adults (blood pressure screening was preserved at around 97% in adults aged 80+, Figure 2C). Blood pressure screening was also maintained in those with a record of care home status (99.2% in March 2019 to 98.7% in March 2023) or learning disability (98.0% in March 2019 to 98.3% in March 2023, Figures 2G and 2H). From December 2022 to March 2023 results indicate an improvement in recorded blood pressure screening across all demographic and clinical subgroups.

**Figure 2.**
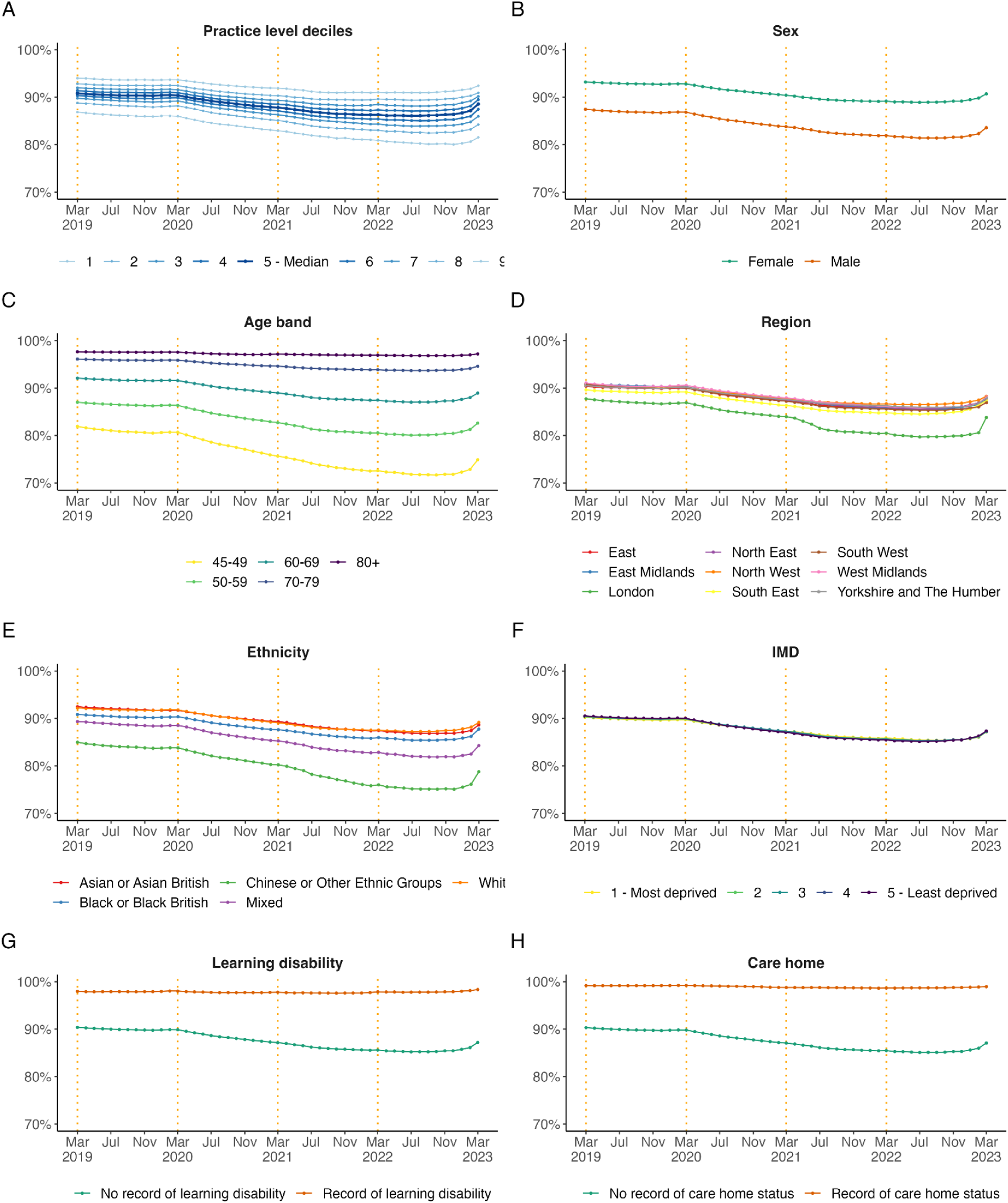
Monthly, unstandardised trends from March 2019 to March 2023 in the percentage of patients aged ≥ 45 with recorded blood pressure in the preceding 5 years in (BP002) broken down by (A) practice level deciles, (B) sex, (C) age band, (D) region, (E) ethnicity, (F) IMD = Indices of Multiple Deprivation, (G) learning disability, and (H) care home status for hypertension. The end of the NHS financial years (March) are highlighted with orange dashed vertical lines..

#### Subgroups for hypertension prevalence (HYP001)

Grouping by demographic subgroups revealed pre-pandemic differences in hypertension recording. In March 2019, the national median by practice was 15.0%, however this was considerably lower in London (10.4%) and in those with an ethnicity record of ‘Mixed’ (6.1%) or ‘Chinese or Other Ethnic Groups’ (6.9%). Hypertension was also less frequently recorded in those living in the most deprived areas (12.8%) compared to those living in the least deprived areas (15.6%). Hypertension was more often recorded in those living in care homes (48.8%) and less frequently in those with learning difficulties (9.6%). The differences observed at pre-pandemic remained similar throughout the study period across most subgroups (Figure 3).

**Figure 3.**
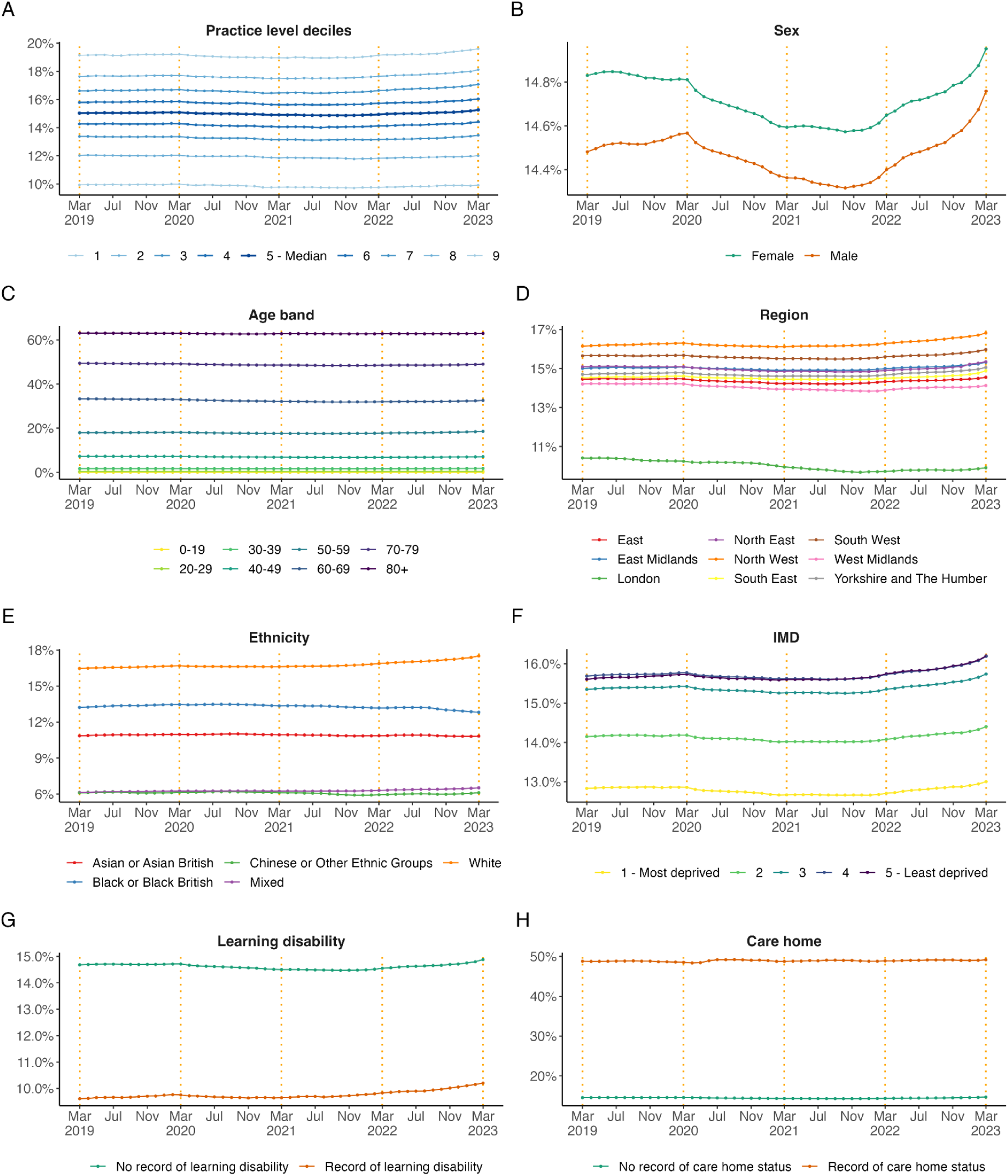
Monthly, unstandardised trends from March 2019 to March 2023 in hypertension prevalence (HYP001) broken down by (A) practice level deciles, (B) sex, (C) age band, (D) region, (E) ethnicity, (F) IMD = Indices of Multiple Deprivation, (G) learning disability, and (H) care home status for hypertension. The end of the NHS financial years (March) are highlighted with orange dashed vertical lines. Note that the range of the y-axis varies by breakdown category to highlight differences between groups.

#### Subgroups for hypertension management in the preceding 12 months in patients aged ≤ 79 (HYP003) and ≥ 80 (HYP007)

Preexisting regional differences between subgroups in blood pressure management in patients diagnosed with hypertension aged ≤ 79 (HYP003) increased during the study period (Figure 4A). In March 2021 the regions South-East and West Midlands had the lowest proportion of patients treated to target, with 39.4% and 41.8% respectively (Figure 5A), compared with the national median by practice of 49.4% and other regions. A similar trend was observed for patients aged ≥ 80 (HYP007, Figures 4B and 5B).

**Figure 4.**
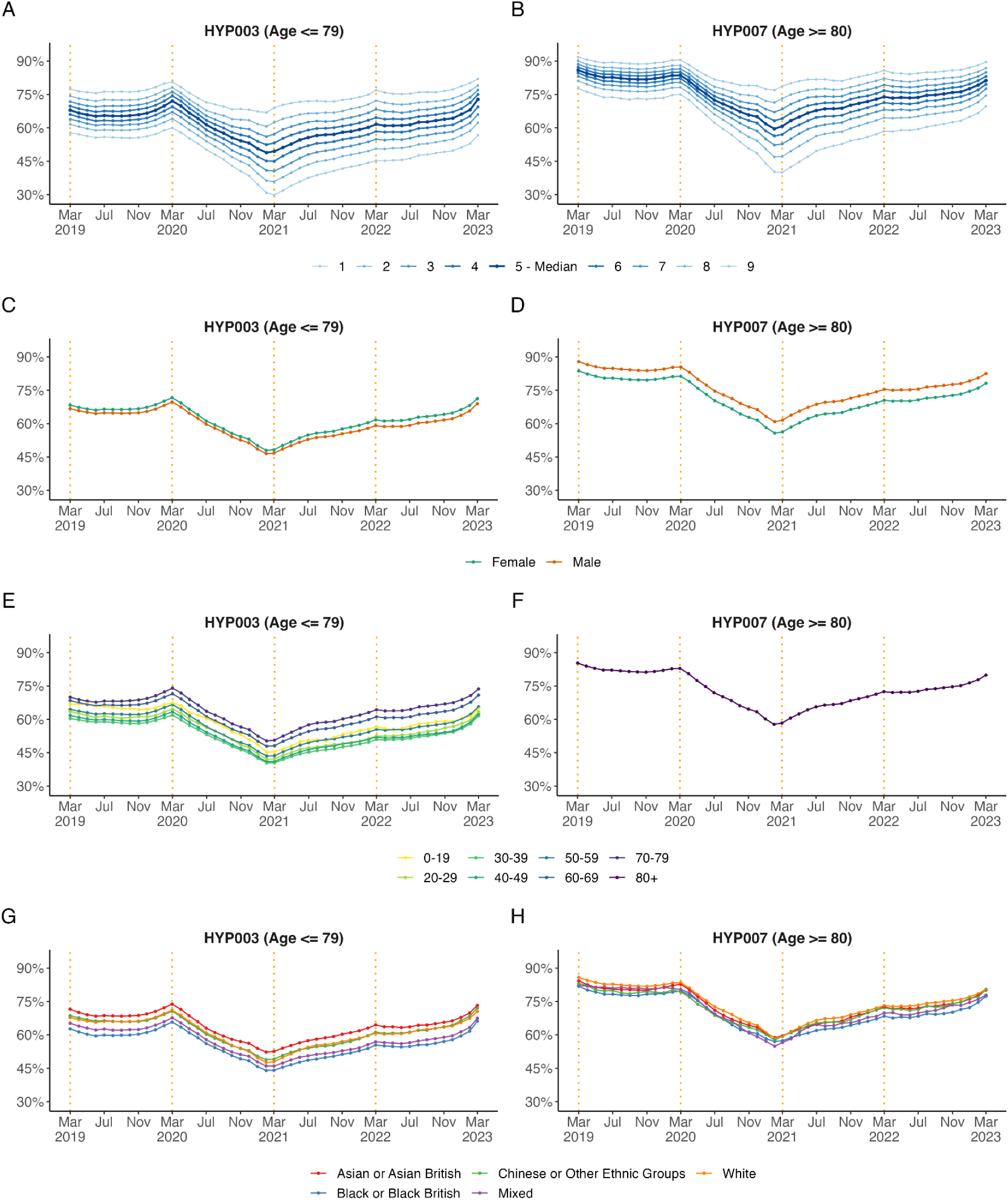
Monthly, unstandardised trends from March 2019 to March 2023 in the percentage of patients diagnosed with hypertension treated to target in the preceding 12 months aged ≤ 79 (HYP003) and ≥ 80 (HYP007) broken down by (A, B) practice level deciles, (C, D) sex, (E, F) age band, (G, H) ethnicity. The end of the NHS financial years (March) are highlighted with orange dashed vertical lines..

**Figure 5.**
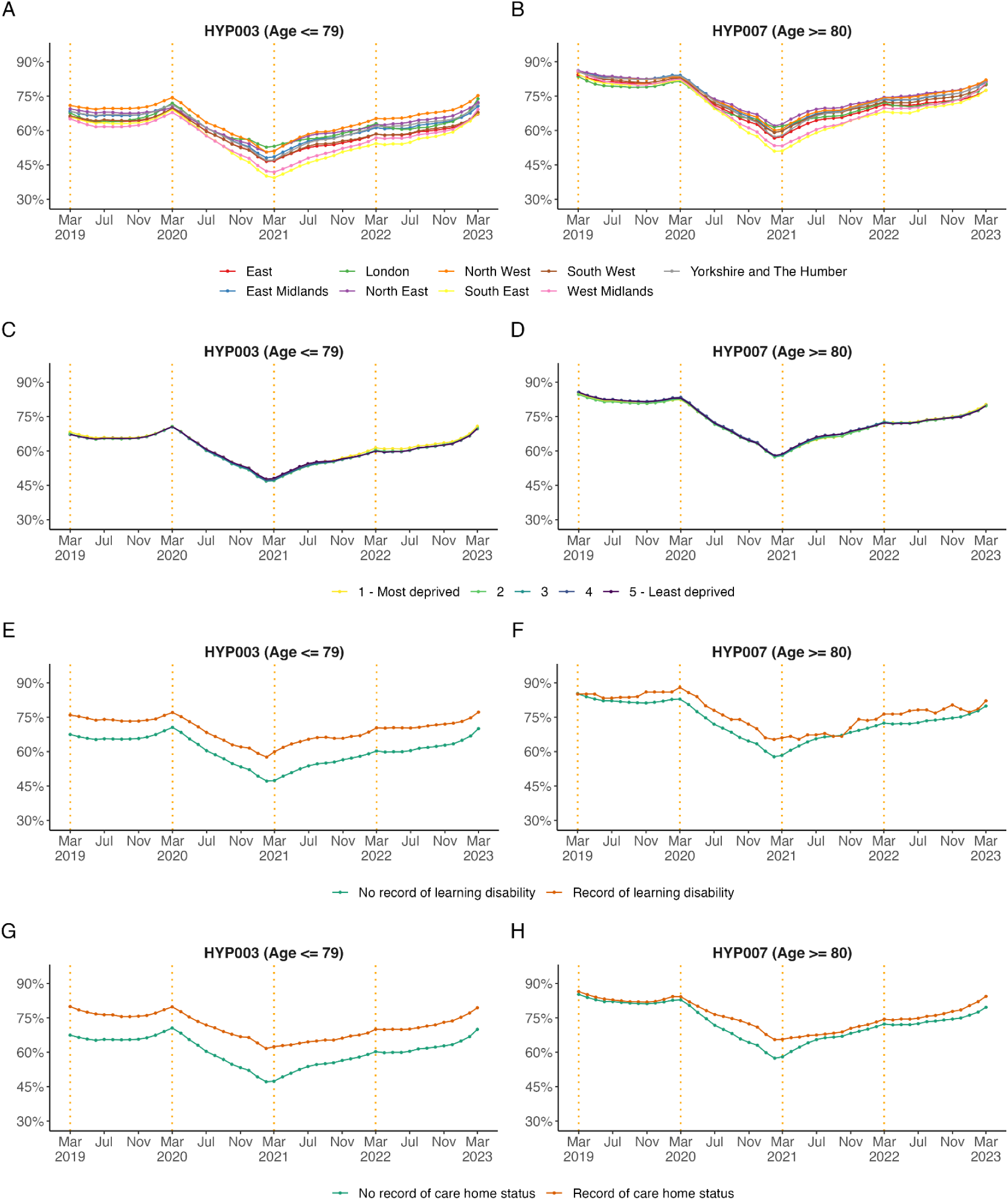
Monthly, unstandardised trends from March 2019 to March 2023 in the percentage of patients diagnosed with hypertension treated to target in the preceding 12 months aged ≤ 79 (HYP003) and ≥ 80 (HYP007) broken down by (A, B) region, (C, D) IMD = Indices of Multiple Deprivation, (E, F) record of learning disability, and (G, H) care home status. The end of the NHS financial years (March) are highlighted with orange dashed vertical lines.

Later in the analysis period, between March 2021 and March 2023, a higher proportion of patients in older age groups (60-69 and 70-79) were reported as having their hypertension treated to target than younger age groups.

Between March 2020 and March 2021 the proportion of patients in care homes with hypertension treated to target reduced by 17.4% in those aged ≤ 79 (HYP003) and by 18.6% in those aged ≥ 80 (HYP007). In March 2023, the proportion of patients with hypertension treated to target had nearly returned to pre-pandemic levels.

## Discussion

### Summary

These results suggested that the pandemic had a substantial impact on the percentage of patients in whom hypertension was classified as not treated to target within the preceding 12 months. Our analyses suggest that this may be attributed to a reduction in blood pressure measurement in the preceding 12 months. We did not observe a substantial impact of the pandemic on the public health QOF standard for blood pressure screening, which is assessed within the preceding 5 years, or on the register for hypertension prevalence.

### Strengths and limitations

This study has a range of strengths. The OpenSAFELY-TPP platform runs analyses across the full raw, pseudonymised, dataset for 25.2 million patients at 2540 practices in England using TPP software. This includes data on codes relevant to the QOF business rules in OpenSAFELY and enables us to report QOF indicators broken down by detailed demographic characteristics and key clinical subgroups. In this study we have reported monthly QOF measures in near real-time.

We acknowledge several limitations in our analyses. Whilst we have included some pre-pandemic data in our analysis, this was limited and therefore we urge caution in interpreting changes in QOF indicators as being caused by the pandemic [30]. Other factors not included in this study may explain some of the changes we observed during our study period. Furthermore, results within regional and demographic subgroups need to be interpreted carefully because we report unstandardised results and did not account for sociodemographic factors (e.g., age differences between subgroups). A recent study found that OpenSAFELY-TPP was largely representative of the general population of England in terms of IMD, age, sex, ethnicity, and causes of death, albeit with relative underrepresentation of practices in London [17].

We have implemented QOF business rules as described in the text description published by NHS Digital, however we would expect our ascertainment of specific patients to deviate from other sources of QOF data for two reasons: (1) as described above OpenSAFELY-TPP has access to the full raw GP record which many not be the case for all other sources of QOF data; and (2) when translating information from the QOF business rules we had to make pragmatic decisions to resolve some ambiguity. For example, all SNOMED-CT codes that could possibly represent a blood pressure recording are provided together by NHS Digital, which required us to manually classify each code as systolic or diastolic to fulfil later QOF business rules and logic (see *findings in context* and Appendix - Table B1).

Finally, we also note that our data will only include clinical codes for blood pressure recording that were carried out in primary care, by a patient at home and correctly captured in a GP system, or in secondary care and returned to GPs as structured data; this may therefore not include results communicated by letter or phone (such as tests requested while a person is in hospital) or indeed blood pressures recorded by patients using home monitoring machines that aren’t forwarded to the GP. However this is a limitation of all EHR analytic work and in line with the methodology already used in the national QOF programme.

### Findings in context

Blood pressure screening and hypertension management are national priorities [31,32] as a modifiable strong risk factor for CVD globally [33]. Hypertension management has been incentivised by QOF for primary care since 2005, and advances in the detection and management of patients with hypertension have been gradually observed [10]. This progression was disrupted by the COVID-19 pandemic. In response to the unparalleled pressure on the health service in the pandemic, some QOF indicators were suspended, including hypertension management in order to support prioritisation of clinical workload [34]. Practices may have rightly deprioritised some care described here to focus on more urgent care needs during the COVID-19 pandemic.

We have reported a prevalence of hypertension that is consistent with the latest available official annual report by NHS Digital covering NHS financial year 2021/22 (see Table E1 in the Appendix) and with the second annual audit from the CVDPREVENT initiative (covering data up to March 2021) in England [13]. Furthermore, our results are consistent with the most recent quarterly data published by CVDPREVENT including data up to June 2022 [35]. A consistent finding across our work, CVDPREVENT, and official QOF publications was the reduction in the proportion of patients with hypertension treated to target in March 2020 and March 2021, however our study reveals continued recovery of care beyond this period, particularly during the first months of 2023. CVDPREVENT also reported a reduction of 21.4% in the proportion of patients recorded as having their blood pressure treated and meeting the NICE guideline target between March 2020 - March 2021 [13]. There are a number of plausible explanations for this observation. The first explanation is a true and concerning increase in the proportion of patients with suboptimal blood pressure control. The second is that those without a blood pressure recording were reported as not having their hypertension treated to target (see Appendix, Figure C2). This finding is likely due to a 42% reduction of recorded blood pressure measurement activity between April 2019 - April 2021 [3]. The third is the apparent prioritisation of blood pressure measurement in those with poorly controlled blood pressure; or fourth that blood pressure measurement was mainly undertaken in those attending GP practices because they were unwell and required an in-person review. Therefore, with services re-prioritised during the pandemic, it is likely that the combination of the second, third and fourth explanation contributed to this observation.

Interestingly, there is a sex and age dependent difference in the proportion of patients with hypertension treated to target. The proportion of female patients with hypertension treated to target was consistently greater than male patients aged ≤ 79 years (Figure 4C). However, in patients ≥ 80 years this is reversed, with a greater proportion of male patients having their hypertension treated to target (Figure 4D). The cause of this trend is beyond the scope of this study but similar patterns have been observed elsewhere [36]. Similarly, the unstandardised results suggest that there is a trend towards a lower proportion of black ethnic patients with hypertension treated to target (Figures 4G and 4H), however this is likely influenced by age differences between ethnicity groups. Variation in hypertension control according to ethnicity has been described widely elsewhere and the cause of which is likely to be multi-faceted [37].

### Implications for policy and future research

The findings of this study suggest that the percentage of patients with hypertension and blood pressure treated to target (HYP003 and HYP007) fell substantially from March 2020 and only partially recovered between February 2021 and March 2022. It is possible that the observed results represent prioritisation of other clinical areas, and the results of this study should not be interpreted as criticism of GPs. The issue has already been recognised and new NHS services were rapidly established in response (e.g. NHS Community Pharmacy Blood Pressure Check Service and BP@home Service) [31,32,38].

In this study, we observed that patients from the most deprived areas had the lowest proportion of patients on the hypertension register (HYP001) and this decreased further during the pandemic (Figure 3F). There was also a trend towards a lower proportion of black ethnic patients with hypertension treated to target, which has also been observed prior to the pandemic in other elements of CVD care [39]. Future research is needed to help understand the reasons for the observed reductions in hypertension management and to further understand the health inequalities to find effective solutions to best address them. The Core20PLUS5, a national NHS England approach to support the reduction of health inequalities at both national and system level, is a new well placed initiative designed to tackle the management of high blood pressure in these important patient groups. We have rapidly developed re-usable analytic code and implementation guidance to support the Core20PLUS5 initiative in OpenSAFELY and beyond [40].

The OpenSAFELY platform could be used for analyses for NHS England, National Institute of Clinical Excellence, and Care Quality Commission on current indicators of clinical care and has the technical ability to prototype new ones. We developed reusable and modifiable analytic code that replicates complex QOF business rules for blood pressure screening, hypertension prevalence, and hypertension management in OpenSAFELY. The additional demographic and clinical data securely accessible through the OpenSAFELY tools also has a number of advantages to current published reports of QOF. It can be used to identify health inequalities amongst regional, demographic or clinical sub-populations (e.g., ethnicity or record of learning disability) in near real-time for new policies and clinical recommendations. Further, all code used in OpenSAFELY is reusable and modifiable and is available under open source licences, an approach recently endorsed in the Data Saves Lives policy paper by the UK [41]. This provides opportunities for the development of code and a more transparent future for operational analysis in the NHS and research using EHR data.

## Conclusion

Although hypertension management indicators were disrupted substantially during the pandemic, this can likely be attributed to a general reduction of blood pressure measurement. Reassuringly, hypertension management indicators have been improving steadily since March 2021 and are now approaching those seen pre-pandemic. Whilst resources were stretched during the pandemic, blood pressure screening was prioritised by general practitioners in older age groups and patients with a record of learning disability or care home status. OpenSAFELY can be used to continuously monitor monthly changes in quality of care indicators to identify significant changes in key clinical subgroups early.

## Administrative

## Data Availability

All data were linked, stored and analysed securely within the OpenSAFELY platform (https://opensafely.org/). Data include pseudonymised data such as coded diagnoses, drugs, and physiological parameters. No free text data were included. All code is shared openly for review and reuse under MIT open license (https://github.com/opensafely/blood-pressure-sro and https://github.com/opensafely/hypertension-sro). Detailed pseudonymised patient data are potentially reidentifiable and therefore not shared.

https://github.com/opensafely/hypertension-sro

## Acknowledgements

We are very grateful for all the support received from the TPP Technical Operations team throughout this work, and for generous assistance from the information governance and database teams at NHS England and the NHS England Transformation Directorate.

## Information governance

NHS England is the data controller for OpenSAFELY-TPP; [TPP is the data processor]; all study authors using OpenSAFELY have the approval of NHS England. This implementation of OpenSAFELY is hosted within the [TPP environment which is] accredited to the ISO 27001 information security standard and is NHS IG Toolkit compliant [42];

Patient data has been pseudonymised for analysis and linkage using industry standard cryptographic hashing techniques; all pseudonymised datasets transmitted for linkage onto OpenSAFELY are encrypted; access to the platform is via a virtual private network (VPN) connection, restricted to a small group of researchers; the researchers hold contracts with

NHS England and only access the platform to initiate database queries and statistical models; all database activity is logged; only aggregate statistical outputs leave the platform environment following best practice for anonymisation of results such as statistical disclosure control for low cell counts [43].

The OpenSAFELY research platform adheres to the obligations of the UK General Data Protection Regulation (GDPR) and the Data Protection Act 2018. In March 2020, the Secretary of State for Health and Social Care used powers under the UK Health Service (Control of Patient Information) Regulations 2002 (COPI) to require organisations to process confidential patient information for the purposes of protecting public health, providing healthcare services to the public and monitoring and managing the COVID-19 outbreak and incidents of exposure; this sets aside the requirement for patient consent [44]. This was extended in November 2022 for the NHS England OpenSAFELY COVID-19 research platform [45]. In some cases of data sharing, the common law duty of confidence is met using, for example, patient consent or support from the Health Research Authority Confidentiality Advisory Group [46].

Taken together, these provide the legal bases to link patient datasets on the OpenSAFELY platform. GP practices, from which the primary care data are obtained, are required to share relevant health information to support the public health response to the pandemic, and have been informed of the OpenSAFELY analytics platform.

## Ethical Approval

This study was approved by the Health Research Authority (REC reference 20/LO/0651) and by the LSHTM Ethics Board (reference 21863).

## Dissemination to participants and related patient and public communities

We will share information and interpretation of our findings through press releases, social media channels, and plain language summary.

## Appendix

### A1. Denominator and numerator rules

**Table A1.** Descriptions of denominator and numerator rules for QOF indicator BP002 (Version 46). Note that the denominator rule 2 and numerator rule 1 select conditions are identical, but describe different actions for cases that do not meet the conditions (next rule vs reject).

|  | Number | Description |
| --- | --- | --- |
| **Denominator** | 1 | Reject patients from the specified population who are aged less than 45 years old. Pass all remaining patients to the next rule. |
|  | 2 | Select patients passed to this rule who had their blood pressure recorded in the 5 year period leading up to and including the payment period end date. Pass all remaining patients to the next rule. |
|  | 3 | Reject patients passed to this rule chose not to have their blood pressure recorded in the 5 year period leading up to and including the payment period end date. Pass all remaining patients to the next rule. |
|  | 4 | Reject patients passed to this rule who registered with the GP practice in the 3 month period leading up to and including the payment period end date. Select the remaining patients. |
| **Numerator** | 1 | Select patients from the denominator who had their blood pressure recorded in the 5 year period leading up to and including the payment period end date. Reject the remaining patients. |

**Table A2.** Descriptions of denominator and numerator rules for QOF indicator HYP_REG / HYP001 (Version 46).

| Number | Description |
| --- | --- |
| 1 | Select patients from the specified population who have a diagnosis of hypertension which has not been subsequently resolved. Reject the remaining patients. |

**Table A3.** Descriptions of denominator and numerator rules for QOF indicator HYP003 (Version 46). Note that the denominator rule 2 and numerator rule 1 select conditions are identical, but describe different actions for cases that do not meet the conditions (next rule vs reject).

|  | Number | Description |
| --- | --- | --- |
| **Denominator** | 1 | Reject patients from the specified population who are aged greater than 79 years old. Pass all remaining patients to the next rule. |
|  | 2 | Select patients passed to this rule who meet all of the criteria below:<br>• Systolic blood pressure value was 140 mmHg or less.<br>• Diastolic blood pressure value was 90 mmHg or less.<br>• Most recent blood pressure recording was in the 12 months leading up to and including the payment period end date.<br>Pass all remaining patients to the next rule. |
|  | 3 | Reject patients passed to this rule who are receiving maximal blood pressure therapy in the 12 months leading up to and including the payment period end date. Pass all remaining patients to the next rule. |
|  | 4 | Reject patients passed to this rule for whom hypertension quality indicator care was unsuitable in the 12 months leading up to and including the payment period end date. Pass all remaining patients to the next rule. |
|  | 5 | Reject patients passed to this rule who chose not to have their blood pressure recorded in the 12 months leading up to and including the payment period end date. Pass all remaining patients to the next rule. |
|  | 6 | Reject patients passed to this rule who chose not to receive hypertension quality indicator care in the 12 months leading up to and including the payment period end date. Pass all remaining patients to the next rule. |
|  | 7 | Reject patients passed to this rule who meet either of the criteria below:<br>• Latest blood pressure reading in the 12 months leading up to and including the payment period end date was above target levels (systolic value of over 140 mmHg and/or a diastolic value of over 90 mmHg), and was followed by two invitations for hypertension monitoring.<br>• Received two invitations for hypertension monitoring and had no blood pressure recordings during the 12 months leading up to and including the achievement date.<br>Pass all remaining patients to the next rule. |
|  | 8 | Reject patients passed to this rule whose earliest hypertension diagnosis was in the 9 months leading up to and including the payment period end date. Pass all remaining patients to the next rule. |
|  | 9 | Reject patients passed to this rule who were recently registered at the practice (patient registered in the 9 month period leading up to and including the payment period end date). Select the remaining patients. |
| **Numerator** | 1 | Select patients from the denominator who meet all of the criteria below:<br>• Systolic blood pressure value was 140 mmHg or less.<br>• Diastolic blood pressure value was 90 mmHg or less.<br>• Most recent blood pressure recording was in the 12 months up to and including the payment period end date.<br>Reject the remaining patients. |

**Table A4.** Descriptions of denominator and numerator rules for QOF indicator HYP007 (Version 46). Note that the denominator rule number 2 and numerator rule number 1 select conditions are identical, but describe different actions for cases that do not meet the conditions (next rule vs reject).

|  | Number | Description |
| --- | --- | --- |
| **Denominator** | 1 | Reject patients from the specified population who are aged less than 80 years old. Pass all remaining patients to the next rule. |
|  | 2 | Select patients passed to this rule who meet all of the criteria below:<br>• Systolic blood pressure value was 150 mmHg or less.<br>• Diastolic blood pressure value was 90 mmHg or less.<br>• Most recent blood pressure recording was in the 12 months leading up to and including the payment period end date.<br>Pass all remaining patients to the next rule. |
|  | 3 | Reject patients passed to this rule who are receiving maximal blood pressure therapy in the 12 months leading up to and including the payment period end date. Pass all remaining patients to the next rule. |
|  | 4 | Reject patients passed to this rule for whom hypertension quality indicator care was unsuitable in the 12 months leading up to and including the payment period end date. Pass all remaining patients to the next rule. |
|  | 5 | Reject patients passed to this rule who chose not to have their blood pressure recorded in the 12 months leading up to and including the payment period end date. Pass all remaining patients to the next rule. |
|  | 6 | Reject patients passed to this rule who chose not to receive hypertension quality indicator care in the 12 months leading up to and including the payment period end date. Pass all remaining patients to the next rule. |
|  | 7 | Reject patients passed to this rule who meet either of the criteria below:<br>• Latest blood pressure reading in the 12 months leading up to and including the payment period end date was above target levels (systolic value of over 150 mmHg and/or a diastolic value of over 90 mmHg), and was followed by two invitations for hypertension monitoring.<br>• Received two invitations for hypertension monitoring and had no blood pressure recordings during the 12 months leading up to and including the achievement date.<br>Pass all remaining patients to the next rule. |
|  | 8 | Reject patients passed to this rule whose earliest hypertension diagnosis was in the 9 months leading up to and including the payment period end date. Pass all remaining patients to the next rule. |
|  | 9 | Reject patients passed to this rule who were recently registered at the practice (patient registered in the 9 month period leading up to and including the payment period end date). Select the remaining patients. |
| **Numerator** | 1 | Select patients from the denominator who meet all of the criteria below:<br>• Systolic blood pressure value was 140 mmHg or less.<br>• Diastolic blood pressure value was 90 mmHg or less.<br>• Most recent blood pressure recording was in the 12 months up to and including the payment period end date.<br>Reject the remaining patients. |

### B1. Codelists

**Table B1.** Names and descriptions of clinical code clusters from the NHS Primary Care Domain Reference Portal used to implement the QOF business rules and further codelists.

| Cluster name | Description |
| --- | --- |
| **Blood pressure** |  |
| BP_COD* | Blood pressure (BP) recording codes |
| BPDEC_COD | Codes indicating the patient has chosen not to have blood pressure procedure |
| **Hypertension** |  |
| BP_COD* | Blood pressure (BP) recording codes |
| BPDEC_COD | Codes indicating the patient has chosen not to have blood pressure procedure |
| HTMAX_COD | Codes for maximal blood pressure (BP) therapy |
| HYP_COD | Hypertension diagnosis codes |
| HYPINVITE_COD | Invite for hypertension care review codes |
| HYPPCADEC_COD | Codes indicating the patient has chosen not to receive hypertension quality indicator care |
| HYPPCAPU_COD | Codes for hypertension quality indicator care unsuitable for patient |
| HYPRES_COD | Hypertension resolved codes |
| **Breakdown variables** |  |
| LD_COD | Learning disability (LD) codes |
| ETHNICITY† | Ethnicity codes |
| CARE_HOME† | NHS care home codes |

*Note.* \* The *BP\_COD* codelist was manually separated into two codelists: one with all codes referring to systolic blood pressure readings and one with all codes referring to diastolic blood pressure.† These codelists were developed and reviewed using the OpenCodelists platform.

### C. Counts of patients in the numerators and denominators of QOF indicators over time

**Figure C1.**
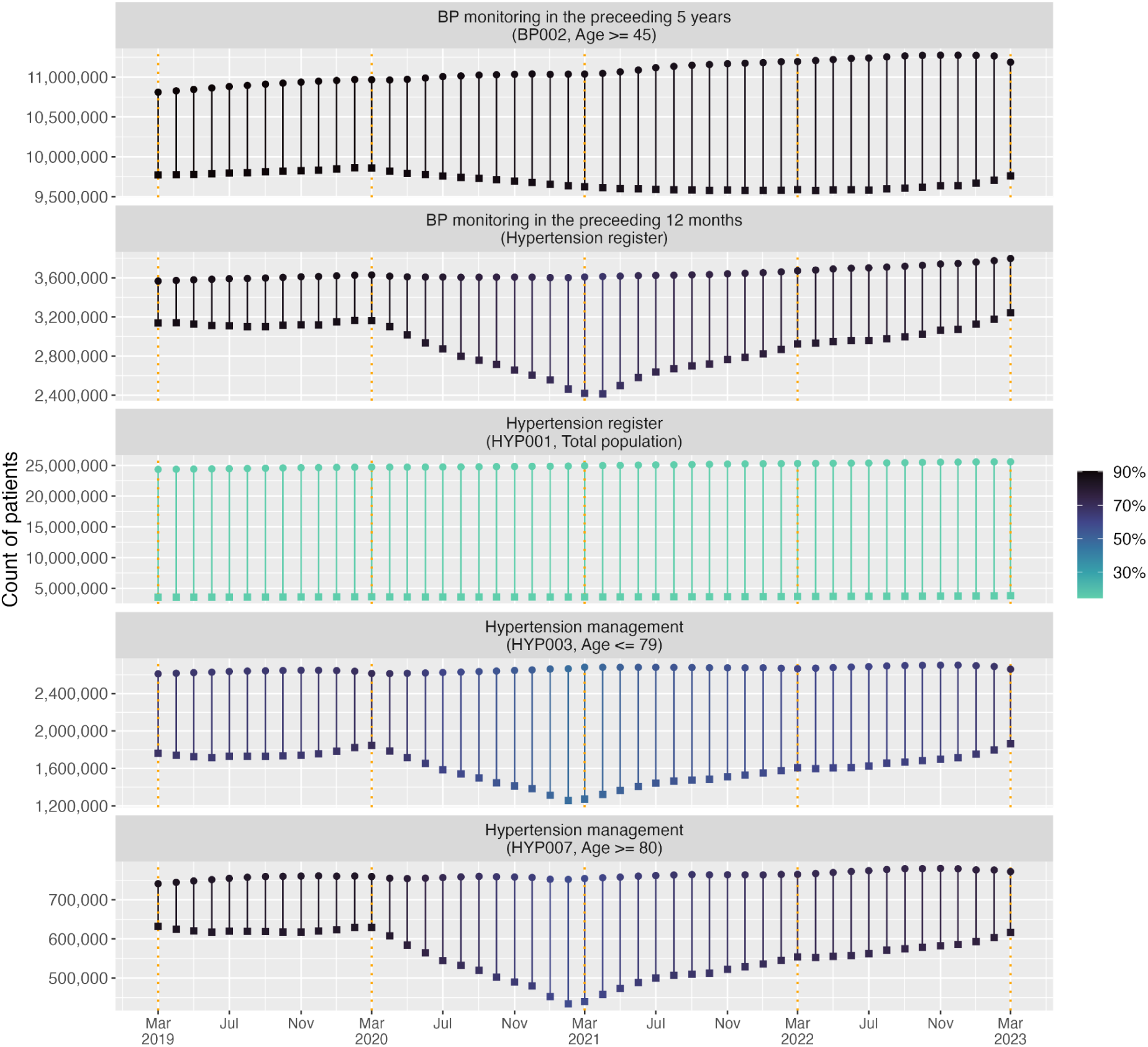
Counts of patients in the numerator and denominator pair for blood pressure and hypertension QOF indicators. The squares indicate the numerator and the circles above represent the denominator. The colour scale indicates the percentage of patients receiving indicated care. The end of the NHS financial years (March) are highlighted with orange dashed vertical lines. Note that the range of the y-axis varies by indicator.

**Figure D1.**
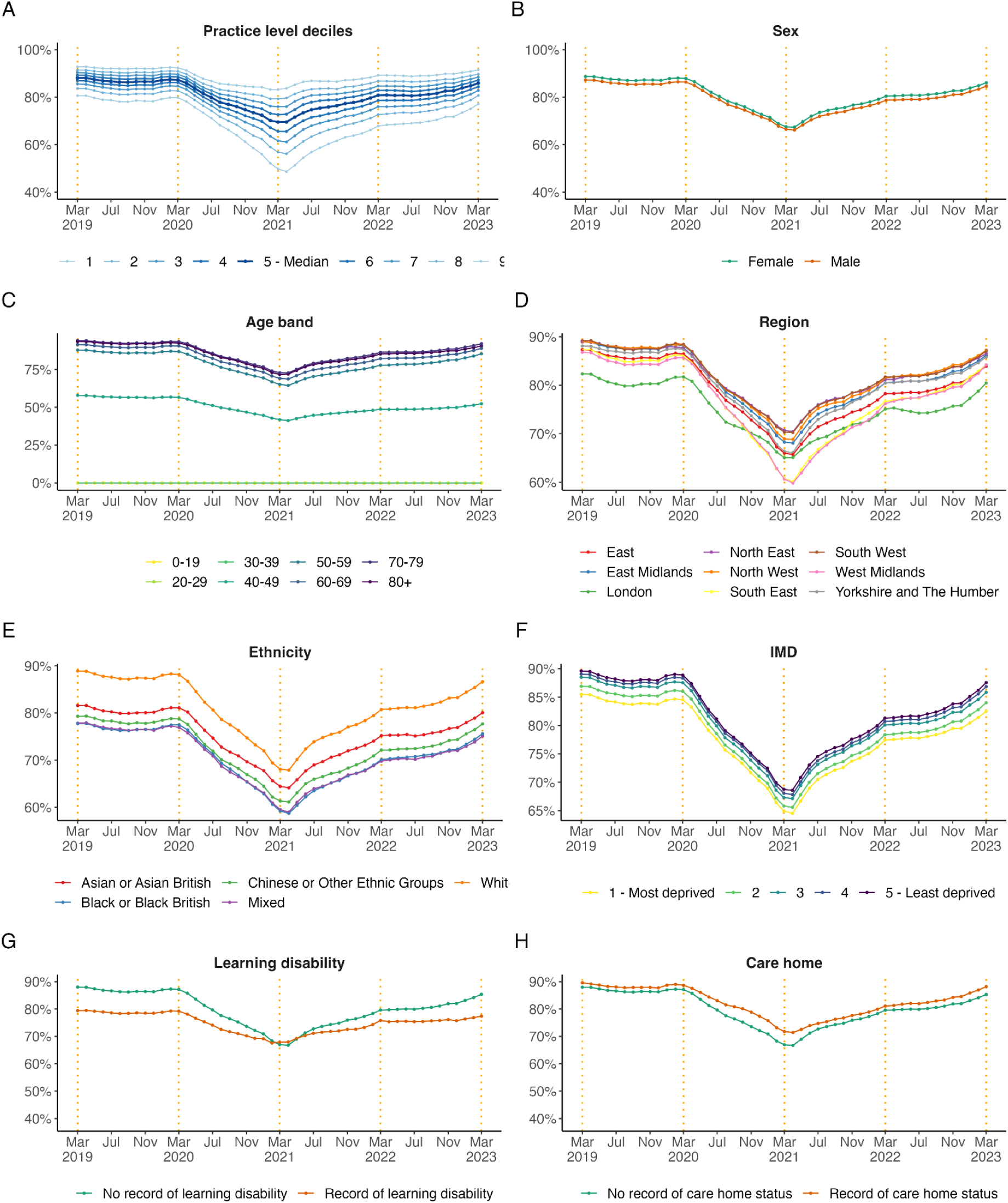
Monthly, unstandardised trends from March 2019 to March 2023 in the percentage of patients with hypertension with recorded blood pressure in the preceding 12 months broken down by (A) practice level deciles, (B) sex, (C) age band, (D) region, (E) ethnicity, (F) IMD = Indices of Multiple Deprivation, (G) learning disability, and (H) care home status for hypertension. The end of the NHS financial years (March) are highlighted with orange dashed vertical lines.

### E. Comparison of QOF results in this study with published results by NHS Digital

**Table E1.**
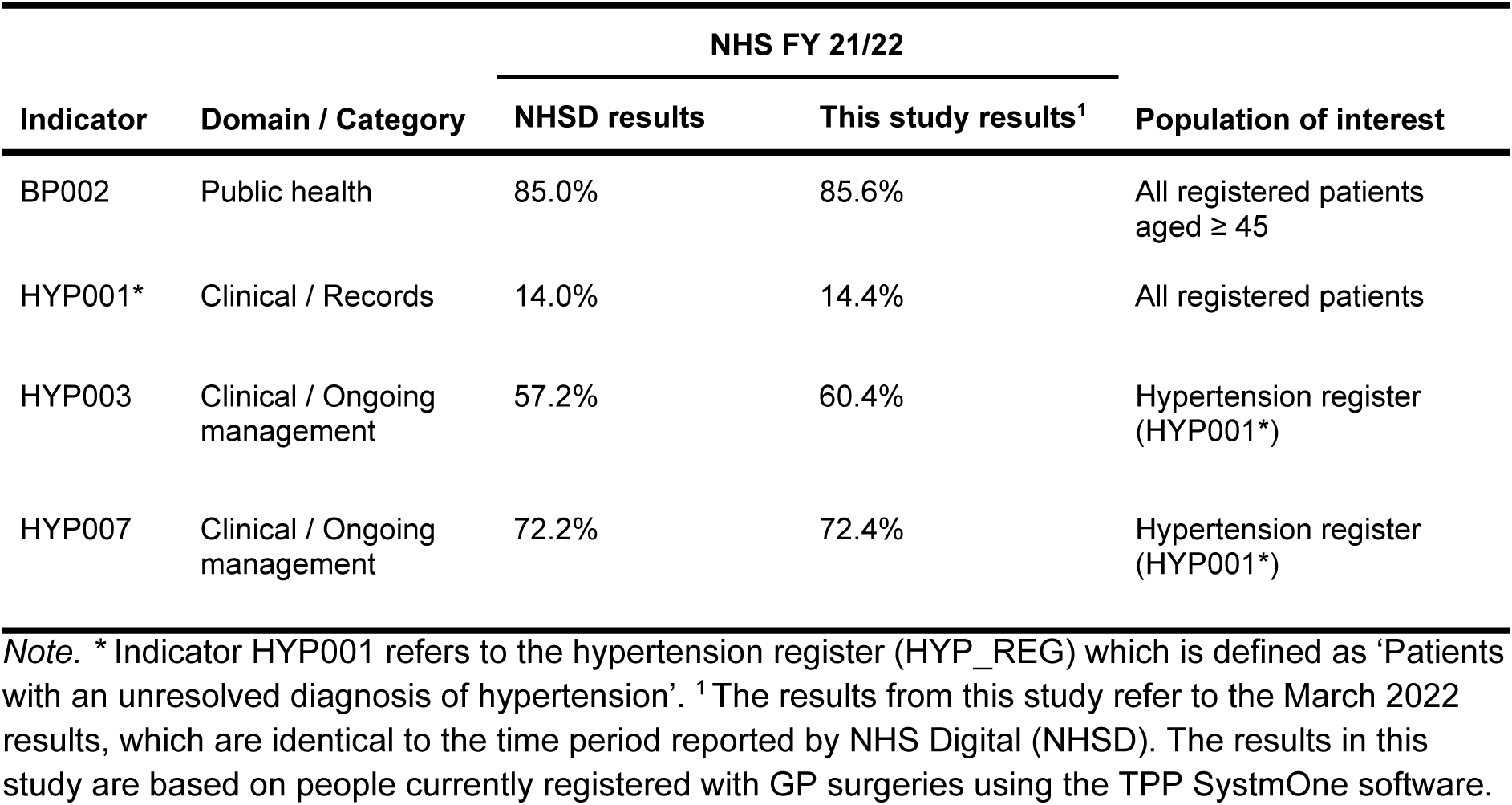
QOF results for the NHS financial year 21/22 in the general population of interest.

| Indicator | Domain / Category | NHS FY 21/22 |  | Population of interest |
| --- | --- | --- | --- | --- |
|  |  | NHSD results | This study results<sup>1</sup> |  |
| BP002 | Public health | 85.0% | 85.6% | All registered patients aged ≥ 45 |
| HYP001* | Clinical / Records | 14.0% | 14.4% | All registered patients |
| HYP003 | Clinical / Ongoing management | 57.2% | 60.4% | Hypertension register (HYP001*) |
| HYP007 | Clinical / Ongoing management | 72.2% | 72.4% | Hypertension register (HYP001*) |

*Note.* \* Indicator HYP001 refers to the hypertension register (HYP\_REG) which is defined as 'Patients with an unresolved diagnosis of hypertension'. <sup>1</sup> The results from this study refer to the March 2022 results, which are identical to the time period reported by NHS Digital (NHSD). The results in this study are based on people currently registered with GP surgeries using the TPP SystemOne software.

## References

1 Moynihan R, Sanders S, Michaleff ZA, et al. Impact of COVID-19 pandemic on utilisation of healthcare services: a systematic review. BMJ Open 2021;11:e045343. doi:10.1136/bmjopen-2020-045343

2 Curtis HJ, MacKenna B, Wiedemann M, et al. OpenSAFELY NHS Service Restoration Observatory 2: changes in primary care activity across six clinical areas during the COVID-19 pandemic. Br J Gen Pract Published Online First: 2023. doi:10.1101/2022.06.01.22275674

3 Fisher L, Curtis HJ, Croker R, et al. Eleven key measures for monitoring general practice clinical activity during COVID-19 using federated analytics on 48 million adults’ primary care records through OpenSAFELY. 2022. doi:10.1101/2022.10.17.22281058

4 Dale CE, Takhar R, Carragher R, et al. The impact of the COVID-19 pandemic on cardiovascular disease prevention and management. Nat Med 2023;29:219–25. doi:10.1038/s41591-022-02158-7

5 Office for National Statistics. Excess deaths in England and Wales. 2022. https://www.ons.gov.uk/peoplepopulationandcommunity/birthsdeathsandmarriages/deaths/articles/excessdeathsinenglandandwales/march2020todecember2021 (accessed 7 Jul 2022).

6 Williamson EJ, Walker AJ, Bhaskaran K, et al. Factors associated with COVID-19-related death using OpenSAFELY. Nature 2020;584:430–6. doi:10.1038/s41586-020-2521-4

7 Silverio A, Di Maio M, Citro R, et al. Cardiovascular risk factors and mortality in hospitalized patients with COVID-19: systematic review and meta-analysis of 45 studies and 18,300 patients. BMC Cardiovasc Disord 2021;21:23. doi:10.1186/s12872-020-01816-3

8 Fuchs FD, Whelton PK. High Blood Pressure and Cardiovascular Disease. Hypertension 2020;75:285–92. doi:10.1161/HYPERTENSIONAHA.119.14240

9 Lim SS, Vos T, Flaxman AD, et al. A comparative risk assessment of burden of disease and injury attributable to 67 risk factors and risk factor clusters in 21 regions, 1990–2010: a systematic analysis for the Global Burden of Disease Study 2010. The Lancet 2012;380:2224–60. doi:10.1016/S0140-6736(12)61766-8

10 Falaschetti E, Mindell J, Knott C, et al. Hypertension management in England: a serial cross-sectional study from 1994 to 2011. The Lancet 2014;383:1912–9. doi:10.1016/S0140-6736(14)60688-7

11 Ashworth M, Medina J, Morgan M. Effect of social deprivation on blood pressure monitoring and control in England: a survey of data from the quality and outcomes framework. BMJ 2008;337:a2030. doi:10.1136/bmj.a2030

12 Xu W, Goldberg SI, Shubina M, et al. Optimal systolic blood pressure target, time to intensification, and time to follow-up in treatment of hypertension: population based retrospective cohort study. BMJ 2015;350:h158. doi:10.1136/bmj.h158

13 Office for Health Improvement & Disparities. CVDPREVENT Second Annual Audit - Using data to drive cardiovascular disease prevention. 2021.

14 NHS Digital. Quality and Outcomes Framework, 2021-22. 2022.https://digital.nhs.uk/data-and-information/publications/statistical/quality-and-outcomes-framework-achievement-prevalence-and-exceptions-data/2021-22 (accessed 10 Nov 2022).

15 National Institute for Health and Care Excellence. Hypertension in adults: diagnosis and management. NICE 2019. https://www.nice.org.uk/guidance/ng136 (accessed 24 Jun 2022).

16 NHS Digital. Quality and Outcomes Framework, 2020-21. 2021.https://digital.nhs.uk/data-and-information/publications/statistical/quality-and-outcomes-framework-achievement-prevalence-and-exceptions-data/2020-21 (accessed 12 Apr 2022).

17 NHS Digital. Quality and Outcomes Framework (QOF) business rules v46.0 2021-2022 baseline release. 2021.https://digital.nhs.uk/data-and-information/data-collections-and-data-sets/data-collections/quality-and-outcomes-framework-qof/quality-and-outcome-framework-qof-business-rules/qof-business-rules-v46.0-2021-2022-baseline-release (accessed 20 Jul 2022).

18 NHS England. JCVI advice in response to the emergence of the B.1.1.529 (Omicron) variant: next steps for deployment. 2022.https://www.england.nhs.uk/coronavirus/wp-content/uploads/sites/52/2021/12/C1468-jvci-advice-in-response-to-the-emergence-of-the-b.1.1.529-omicron-variant-next-steps-for-deployment.pdf (accessed 16 Aug 2022).

19 NHS England and NHS Improvement. Update on Quality Outcomes Framework changes for 2022/23. 2022.https://www.england.nhs.uk/gp/investment/gp-contract/quality-on-outcomes-framework-qof-changes-for-2022-23-and-qof-guidance/

20 Green ACA, Curtis HJ, Higgins R, et al. Trends, variation, and clinical characteristics of recipients of antiviral drugs and neutralising monoclonal antibodies for covid-19 in community settings: retrospective, descriptive cohort study of 23.4 million people in OpenSAFELY. BMJ Med 2023;2. doi:10.1136/bmjmed-2022-000276

21 Schultze A, Nightingale E, Evans D, et al. Mortality among Care Home Residents in England during the first and second waves of the COVID-19 pandemic: an observational study of 4.3 million adults over the age of 65. Lancet Reg Health – Eur 2022;14. doi:10.1016/j.lanepe.2021.100295

22 The OpenSAFELY Collaborative, Curtis HJ, Inglesby P, et al. Trends and clinical characteristics of COVID-19 vaccine recipients: a federated analysis of 57.9 million patients’ primary care records in situ using OpenSAFELY. 2021. doi:10.1101/2021.01.25.21250356

23 The OpenSAFELY Collaborative, Curtis HJ, MacKenna B, et al. OpenSAFELY: impact of national guidance on switching anticoagulant therapy during COVID-19 pandemic. Open Heart 2021;8:e001784. doi:10.1136/openhrt-2021-001784

24 Fisher L, Hopcroft LEM, Rodgers S, et al. Changes in English medication safety indicators throughout the COVID-19 pandemic: a federated analysis of 57 million patients’ primary care records in situ using OpenSAFELY. 2022. doi:10.1101/2022.05.05.22273234

25 Andrews C, Schultze A, Curtis H, et al. OpenSAFELY: Representativeness of electronic health record platform OpenSAFELY-TPP data compared to the population of England. Wellcome Open Res 2022;7:191. doi:10.12688/wellcomeopenres.18010.1

26 NHS. The NHS Long Term Plan. 2019. https://www.longtermplan.nhs.uk/wp-content/uploads/2019/08/nhs-long-term-plan-version-1.2.pdf (accessed 28 Jul 2021).

27 Wickham H. ggplot2: Elegant graphics for data analysis. Springer-Verlag New York 2016. https://ggplot2.tidyverse.org

28 Iannone R, Cheng J, Schloerke B. gt: Easily create presentation-ready display tables. 2022. https://CRAN.R-project.org/package=gt

29 Oswald M, Laverty L. Data Sharing in a Pandemic: Three Citizens’ Juries – Juries Report. 2021. https://arc-gm.nihr.ac.uk/media/Resources/ARC/Digital%20Health/Citizen%20Juries/12621_NIHR_Juries_Report_ELECTRONIC.pdf

30 Walker AJ, Croker R, Curtis HJ, et al. Trends in antidepressant prescribing in England. Lancet Psychiatry 2021;8:278–9. doi:10.1016/S2215-0366(21)00081-X

31 NHS England. Case study: NHS community pharmacy blood pressure check service. 2021. https://web.archive.org/web/20220707071627/https://www.england.nhs.uk/primary-care/pharmacy/pharmacy-integration-fund/pharmacy-integration-fund-case-studies/community-pharmacy-blood-pressure-check-service/ (accessed 7 Jul 2022).

32 NHS England. NHS community pharmacy hypertension case-finding advanced service (NHS Community Pharmacy Blood Pressure Check Service). 2021. https://www.england.nhs.uk/wp-content/uploads/2021/11/B0953-NHS-community-pharmacy-blood-pressure-check-service-specification.pdf (accessed 2 Jul 2022).

33 Yusuf S, Joseph P, Rangarajan S, et al. Modifiable risk factors, cardiovascular disease, and mortality in 155 722 individuals from 21 high-income, middle-income, and low-income countries (PURE): a prospective cohort study. The Lancet 2020;395:795–808. doi:10.1016/S0140-6736(19)32008-2

34 NHS England. Temporary GP contract changes to support COVID-19 vaccination programme. 2021.https://www.england.nhs.uk/coronavirus/documents/temporary-gp-contract-changes-to-support-covid-19-vaccination-programme/ (accessed 19 Jul 2022).

35 NHS Benchmarking Network. CVDPREVENT Quarterly reporting now available. NHS Benchmarking Netw. 2022.https://www.nhsbenchmarking.nhs.uk/news/cvdprevent-quarterly-reporting-coming-soon (accessed 25 Jan 2023).

36 NHS England. Report of the Review of the Quality and Outcomes Framework in England. 2018. https://www.england.nhs.uk/wp-content/uploads/2018/07/quality-outcome-framework-report-of-the-review.pdf (accessed 19 Jul 2022).

37 Marshall IJ, Wolfe CDA, McKevitt C. Lay perspectives on hypertension and drug adherence: systematic review of qualitative research. BMJ 2012;345:e3953. doi:10.1136/bmj.e3953

38 NHS England. Home blood pressure monitoring. 2021.https://web.archive.org/web/20230417064030/https://www.england.nhs.uk/ourwork/clinical-policy/cvd/home-blood-pressure-monitoring/ (accessed 15 Jun 2023).

39 Eastwood SV, Mathur R, Sattar N, et al. Ethnic differences in guideline-indicated statin initiation for people with type 2 diabetes in UK primary care, 2006–2019: A cohort study. PLOS Med 2021;18:e1003672. doi:10.1371/journal.pmed.1003672

40 Brown A. Generating data on the NHS England Core20PLUS5 inequality groups using OpenSAFELY in GP records. 2023.https://www.bennett.ox.ac.uk/blog/2023/01/generating-data-on-the-nhs-england-core20plus5-inequality-groups-using-opensafely-in-gp-records/ (accessed 8 Mar 2023).

41 Department of Health and Social Care. Data saves lives: reshaping health and social care with data. 2022.https://www.gov.uk/government/publications/data-saves-lives-reshaping-health-and-social-care-with-data/data-saves-lives-reshaping-health-and-social-care-with-data (accessed 30 Jun 2022).

42 NHS Digital. Data Security and Protection Toolkit. 2020.https://digital.nhs.uk/data-and-information/looking-after-information/data-security-and-information-governance/data-security-and-protection-toolkit (accessed 30 Apr 2020).

43 NHS Digital. ISB1523: Anonymisation Standard for Publishing Health and Social Care Data. https://digital.nhs.uk/data-and-information/information-standards/information-standards-and-data-collections-including-extractions/publications-and-notifications/standards-and-collections/isb1523-anonymisation-standard-for-publishing-health-and-social-care-data (accessed 30 Apr 2020).

44 Secretary of State for Health and Social Care - UK Government. Coronavirus (COVID-19): notification to organisations to share information. 2020.https://web.archive.org/web/20221101161400/https://www.gov.uk/government/publications/covid-19-notification-to-gps-and-nhs-england-to-share-information (accessed 30 Jan 2022).

45 Secretary of State for Health and Social Care - UK Government. Coronavirus (COVID-19): notification to organisations to share information. 2022.https://www.gov.uk/government/publications/coronavirus-covid-19-notification-to-organisations-to-share-information/coronavirus-covid-19-notice-under-regulation-34-of-the-health-service-control-of-patient-information-regulations-2002

46 NHS Health Research Authority. Confidentiality Advisory Group. https://www.hra.nhs.uk/about-us/committees-and-services/confidentiality-advisory-group/ (accessed 30 Jan 2023).

